# Durability of protection post-primary COVID-19 vaccination in the US: matched case-control study

**DOI:** 10.1101/2022.01.05.22268648

**Authors:** Amanda Zheutlin, Miles Ott, Ran Sun, Natalia Zemlianskaia, Meagan Rubel, Jennifer Hayden, Breno Neri, Tripthi Kamath, Najat Khan, Sebastian Schneeweiss, Khaled Sarsour

**Affiliations:** Data Sciences, Research & Development, Janssen Pharmaceuticals; Titusville, NJ; Data Sciences, Research & Development, Janssen Pharmaceuticals; Cambridge, MA; Data Sciences, Research & Development, Janssen Pharmaceuticals; South San Francisco, CA; Data Sciences, Research & Development, Janssen Pharmaceuticals; San Diego, CA; Division of Pharmacoepidemiology, Department of Medicine, Brigham and Women’s Hospital and Harvard Medical School; Boston, MA

## Abstract

**Background:** Intrinsic durability of immune responses elicited by COVID-19 vaccines will drive vaccine effectiveness long-term across settings and may differ by vaccine type. We aimed here to determine durability of protection of three COVID-19 vaccines BNT162b2, mRNA-1273 and Ad26.COV2.S following primary vaccination against breakthrough infections, hospitalisations, and intensive care unit (ICU) admissions in the United States (US).

**Methods:** Using national claims and laboratory data covering 168 million lives, we conducted a matched case-control study with fully vaccinated individuals between January 1 and September 7, 2021. Odds ratios (OR) for developing outcomes in months two through six following primary vaccination were estimated relative to the first month after primary vaccination for each vaccine separately. Results compare each vaccine to itself and are not directly comparative. Odds ratios were translated into vaccine effectiveness (VE) using assumptions about event rates in an unvaccinated cohort.

**Findings:** Relative to its baseline, stable protection was observed for the single-shot Ad26.COV2.S against infections and severe disease. Relative to their baseline protection waned overtime against infections for BNT162b2 and mRNA-1273 and against hospitalisations for BNT162b2. No waning of baseline protection was observed at any time for ICU admissions for all three vaccines. Calculated baseline VE was consistent with the published literature.

**Interpretation:** While the starting protection level provided by the primary series may differ by vaccine type and mechanism of action, this study demonstrated by comparing each vaccine to its own baseline protection that the three vaccines in three separate populations may have different durability profiles. Further investigation is required to fully characterize the durability profile of the three vaccines. Moreover, as the COVID-19 pandemic continues, and as more countries and populations implement a standard of care consisting of three doses of the mRNA vaccines or two doses of Ad26.COV2.S, further investigation is critical to understand the level of protection and the durability of response over longer periods, novel variants and in response to homologous and heterologous boosting.

## Main Text

Clinical trials have demonstrated high vaccine efficacy of the primary series in protection against coronavirus disease 2019 (COVID-19) for all three US-authorized or approved vaccines (Ad26.COV2.S, BNT162b2, and mRNA-1273) (*1–3*), and subsequent studies have confirmed high vaccine effectiveness (VE) against infection and severe disease in the real-world (*4–8*). Preliminary data on waning VE and the emergence of severe acute respiratory syndrome coronavirus 2 (SARS-CoV-2) variants of concern have prompted regulatory approval or emergency use authorization for booster doses in the United States for all adults. Subsequently, the CDC recommended that BNT162b2 or mRNA-1273 are preferred to Ad26.COV2.S.

Studies on the durability of all three US vaccines in the general population have been limited to date, with many focusing on high-risk populations or specific geographic areas (*7, 9–13*). Two distinct factors may contribute to the durability of protection or declining effectiveness of a given vaccine. One is the waning immunity against a stable pathogen, due to declines in antibody levels, B-cell memory, and/or T-cell populations. Second is the emergence of novel variants of the pathogen. Without access to pathogen strain data and repeated measures of immune responses, large population-based observational studies may not be able to directly disentangle the effects of these two factors. Nonetheless, there is still a need to understand the overall durability of vaccine protection among the general adult population. Moreover, durability of responses to primary vaccine regimens remains especially important for the management of the pandemic globally, as much of the world remains unvaccinated or only recently vaccinated with the primary series. As such, it is critical to assess the degree to which protection is sustained for each vaccine over time in the general adult population. In this study, we report the durability of protection against COVID-19 in over 17 million vaccinated individuals following primary vaccination for each approved or authorized vaccine against breakthrough infections, hospitalizations, and intensive care unit (ICU) admissions, while accounting for calendar time, residential location, age, sex, and comorbid conditions.

## Materials and Methods

### Study design

We used a matched case-control design conducted with three cohorts of vaccinated individuals to assess the durability of vaccine-induced protection against SARS-CoV-2 infections, COVID-19-related hospitalizations, and COVID-19-related ICU admissions in the United States between January 1 and September 7, 2021, as per Ray and Klein (*26, 27*). Since the three vaccines were authorized at different times resulting in differences in both vaccination and follow-up time, as well as demographics (e.g., older individuals were eligible for vaccination prior to authorization of Ad26.COV2.S), we analyzed the mRNA vaccine doses in two different ways. We required the second mRNA vaccine dose to be on or after February 27, 2021, the first date at which all three vaccines were available. We also analyzed all available data where a first mRNA dose was on or after January 1, 2021. For Ad26.COV2.S, we required the first (and only) dose to be on or after February 27, 2021 (Fig. S6).

We compared the odds of breakthrough infection, hospitalization, or ICU admission in each subsequent 28-day window (months 2 through 6+ since vaccination) relative to the first month since vaccination (reference month lasting 28 days). Here, durability was defined as constant odds for the outcome of interest across time. Follow-up started after a 21-day period following the final vaccination dose; day 22 is thus the beginning of the first month of follow-up (Fig. S6). We selected this timeframe to allow for the 14 days required for full vaccination and an additional 7-day washout period to ensure COVID-19-related outcomes included in our study were limited to those due to COVID-19 infections acquired *after* completion of the primary vaccination (*28*). Since the CDC guidelines recommend waiting 5 to 7 days after exposure before being tested (*29*), test results and hospitalizations recorded soon after vaccination (i.e., 15–21 days after the last dose) may reflect infections and exposures occurring prior to vaccination.

The collection and analysis of these data did not qualify as human subjects research under the Common Rule and were not subject to institutional review board review (*30*). The New England Institutional Review Board approved this exemption (no.1-9757-1). Upon reasonable request, researchers may retrieve access to the data and analytics infrastructure for prespecified collaborative analyses.

### Data

We analyzed de-identified person-level longitudinal data captured from medical and pharmacy claims, laboratory tests and results, and hospital chargemaster data from over 70 sources in the United States aggregated by HealthVerity. This database has been used in other COVID-19 research studies (*31–34*). We drew cohorts for the current study from a broadly defined COVID-19 dataset where individuals were included based on having any documentation of COVID-19 related diagnoses, laboratory orders, procedures, treatments, or COVID-19 vaccinations. The dataset contained all major insurance types in the United States in proportions representative of the US population: Medicaid (19%), Medicare (14%), commercial (60%), and federal (5%) (*15*). Race and ethnicity information was not available as they were considered personally identifiable information by HealthVerity.

For the more than 168 million individuals included in the source data, all available health data regardless of relation to COVID-19 were included. Both open (claims sent from providers to payers) and closed (adjudicated) claims were used. While open claims do not include participant enrollment (limiting certainty of observability), they capture events closer to real-time since there is no lag caused by the adjudication process. To avoid inclusion of persons whose care would not be continuously available in this database (e.g., persons that do not routinely receive medical care from an included healthcare system), we required prior utilization in the two years preceding vaccination for inclusion (see Population section below).

### Population

We defined eligible individuals as adults aged 18 years or older who were vaccinated following the recommended primary vaccination schedule. For mRNA-1273, two doses 28 to 42 days apart were required; for BNT162b2, two doses 21 to 42 days apart were required; and for Ad26.COV2.S, one dose was required (*35*). For cohort analyses using a subset of data available from February 27, 2021 to September 7, 2021, we required the second dose of mRNA-1273 or BNT162b2 and the first dose of Ad26.COV2.S to be on or after February 27, 2021, the earliest point when all three were available. For full cohorts, we included individuals with a first dose of mRNA-1273 or BNT162b2 on January 1, 2021, or later. Cohort entry began 21 days after the last vaccine dose (Fig. S6).

We excluded individuals with 1) any positive diagnostic or antibody laboratory test or a diagnostic ICD-10 code for COVID-19 before cohort entry; 2) missing age, sex, or geographic information (three-digit zip code); 3) heterologous primary series of COVID-19 vaccines (e.g., one dose of BNT162b2 followed by one dose of mRNA-1273, or vice versa); 4) more or fewer than the required number of doses (e.g., a second dose of the Ad26.COV2.S vaccine); and 5) no medical claims in the 2 years before their last required vaccine dose to ensure observability of patient information.

### Exposure

The exposure of interest was the time in months from cohort entry to the outcome of interest. We grouped follow-up time (occurring between 21-days post completed vaccination and September 7, 2021) into five exposure categories: month 1 or reference month (1–28 days after follow-up started), month 2 (29–56 days), month 3 (57–84 days), month 4 (85–112 days), month 5 (113–140 days), and month 6+ (141 or more days).

### Cases and controls

Within each of the three separate vaccine cohorts, cases were those who experienced the outcome of interest while controls were those from the same vaccine cohort who did not experience the outcome of interest. We matched each case to up to 10 controls who had entered the cohort prior to or on the calendar date of the case outcome. Cases and controls were matched on calendar time, location (three-digit zip code), sex, age (5-year category), and Gagne comorbidity score category (<2, 2–3, 4–5, 6+) using risk set sampling without replacement (Fig. S6) (*36, 37*). The first outcome of interest was infection identified by either a COVID-19 ICD-10-CM diagnosis code (U07.1, U07.2) or a positive polymerase chain reaction (PCR) laboratory test. The case date was defined as the date of the first occurrence of a relevant code or test result during the study period.

We included incident hospitalization and ICU admission as additional outcomes to index more severe disease. A COVID-19-related hospitalization was defined as an inpatient hospital encounter during which a COVID-19 ICD-10-CM code (U07.1, U07.2) or positive PCR laboratory test result was recorded. For case events, we required the initial admission date to be during our study period to further avoid hospitalizations due to infections acquired prior to full vaccination. COVID-19-related ICU admissions were defined as an inpatient hospital encounter as described above that additionally included admission to an ICU. For both outcomes, the first of this type of event during the study period was considered the case event and the case event date was set as the hospital admission date. Controls were selected and matched as described above for breakthrough infections.

### Statistical analysis

To assess the durability of protection, we fitted conditional logistic regression models separately for each vaccine and outcome combination, resulting in nine models. We fitted conditional logistic-regression models to estimate the effect of each additional month after receipt of the primary series vaccine dose on the odds of the outcome of interest. The models estimated the effects of time measured in month increments using month 1 as the reference category. ORs with 95% confidence intervals (OR; 95% CI) measured change in baseline protection relative to the referent month.

As ORs here reflect change in risk over time from potentially different baseline groups, we translated ORs for infection and hospitalization to vaccine effectiveness (VE) by incorporating all eligible vaccinated individuals (not restricted to case-control matched groups) and unvaccinated individuals. Sensitivity analyses stratifying by age and comorbidity score were also conducted (see Supplementary Methods below).

### Translating ORs to VE

We estimated corresponding VE for each of the three vaccines (*38–40*) for COVID-19 infection and COVID-19-related hospitalization outcomes. All eligible vaccinated and unvaccinated individuals were used for these estimates. Vaccinated individuals were eligible if they met the inclusion and exclusion criteria described in the main analysis. Unvaccinated individuals were defined as those without any record of any vaccine and were excluded if they met criteria 1, 2, or 5, as described for the vaccinated cohorts in the main text. Cohort entry for unvaccinated individuals used in primary vaccine regimens using a subset of data available from February 27, 2021, to September 7, 2021, was also conducted, as that was the earliest possible cohort entry date for any vaccinated individual. For the full mRNA cohorts, cohort entry for the unvaccinated was set to February 14, 2021. For criterion 5, prior medical claims were required in the 2 years preceding cohort entry.

To estimate VE, we combined the OR for the outcomes for each of the three vaccines from the conditional logistic regression models described in the main manuscript (Table 3) with models to estimate outcome-specific baseline odds among all vaccinated individuals for each vaccine.

**Table 1.** Demographic characteristics of vaccine recipients for three vaccine cohorts.

|  | **Ad26.COV2.S**<br>**n (%)** | **All BNT162b2**<br>**n (%)** | **BNT162b2**<br>**n (%)**<br>(for primary vaccine<br>regimens using a subset<br>of data available from<br>February 27, 2021 –<br>September 7, 2021) | **All mRNA-1273**<br>**n (%)** | **mRNA-1273**<br>**n (%)**<br>(for primary vaccine<br>regimens using a subset<br>of data available from<br>February 27, 2021 –<br>September 7, 2021) |
| --- | --- | --- | --- | --- | --- |
| N | 1,761,498 | 7,903,410 | 6,700,042 | 7,352,527 | 6,466,763 |
| Age group<br>(years) |  |  |  |  |  |
| 18–24 | 156,961 (8.9%) | 683,915 (8.7%) | 651,453 (9.7%) | 457,258 (6.2%) | 435,240 (6.7%) |
| 25–29 | 98,955 (5.6%) | 451,780 (5.7%) | 421,484 (6.3%) | 333,920 (4.5%) | 309,484 (4.8%) |
| 30–34 | 111,886 (6.4%) | 512,443 (6.5%) | 478,469 (7.1%) | 379,781 (5.2%) | 351,542 (5.4%) |
| 35–39 | 123,369 (7.0%) | 537,280 (6.8%) | 499,979 (7.5%) | 407,096 (5.5%) | 374,108 (5.8%) |
| 40–44 | 130,393 (7.4%) | 544,381 (6.9%) | 504,248 (7.5%) | 422,532 (5.7%) | 386,331 (6.0%) |
| 45–49 | 144,644 (8.2%) | 573,498 (7.3%) | 529,015 (7.9%) | 451,982 (6.1%) | 414,420 (6.4%) |
| 50–54 | 181,413 (10.3%) | 681,287 (8.6%) | 625,690 (9.3%) | 569,539 (7.7%) | 526,020 (8.1%) |
| 55–59 | 213,464 (12.1%) | 797,898 (10.1%) | 727,649 (10.9%) | 699,247 (9.5%) | 648,792 (10.0%) |
| 60–64 | 218,591 (12.4%) | 869,285 (11.0%) | 787,033 (11.7%) | 813,055 (11.1%) | 753,929 (11.7%) |
| 65–69 | 143,103 (8.1%) | 662,119 (8.4%) | 549,080 (8.2%) | 907,660 (12.3%) | 799,975 (12.4%) |
| 70–74 | 104,923 (6.0%) | 553,754 (7.0%) | 408,479 (6.1%) | 766,263 (10.4%) | 642,708 (9.9%) |
| 75–79 | 62,574 (3.6%) | 371,754 (4.7%) | 235,230 (3.5%) | 485,138 (6.6%) | 374,793 (5.8%) |
| 80–84 | 32,566 (1.8%) | 230,994 (2.9%) | 127,075 (1.9%) | 274,499 (3.7%) | 202,833 (3.1%) |
| 85+ | 38,455 (2.2%) | 432,561 (5.5%) | 154,822 (2.3%) | 383,911 (5.2%) | 246,018 (3.8%) |
| Sex |  |  |  |  |  |
| Male | 845,728 (48.0%) | 3,286,709 (41.6%) | 2,869,428 (42.8%) | 3,125,010 (42.5%) | 2,794,551 (43.2%) |
| Female | 915,727 (52.0%) | 4,616,322 (58.4%) | 3,830,239 (57.2%) | 4,227,255 (57.5%) | 3,671,984 (56.8%) |
| Comorbidity<br>score category |  |  |  |  |  |
| <2 | 1,576,908 (89.5%) | 6,865,542 (86.9%) | 5,993,827 (89.4%) | 6,375,144 (86.7%) | 5,665,073 (87.6%) |
| 2–3 | 126,114 (7.2%) | 662,133 (8.4%) | 483,613 (7.2%) | 635,831 (8.6%) | 533,955 (8.3%) |
| 4–5 | 34,213 (1.9%) | 212,538 (2.7%) | 129,349 (1.9%) | 199,039 (2.7%) | 157,664 (2.4%) |
| 6+ | 24,263 (1.4%) | 163,197 (2.1%) | 93,253 (1.4%) | 142,513 (1.9%) | 110,071 (1.7%) |
| Medicaid | 154,072 (8.7%) | 570,587 (7.2%) | 485,282 (7.2%) | 554,110 (7.5%) | 475,092 (7.3%) |
| insured |  |  |  |  |  |

**Table 2.** Age and comorbidity scores of vaccine recipients for three vaccine cohorts.

| **Vaccine** | **Age,**<br>**mean (SD)** | **Gagne combined**<br>**comorbidity score,**<br>**mean (SD)** | **Gagne combined**<br>**comorbidity score,**<br>**Median (IQR)** |
| --- | --- | --- | --- |
| Ad26.COV2.S | 50.9 (17.5) | 0.41 (1.28) | 0.0 (0.0) |
| All BNT162b2 | 53.1 (19.4) | 0.53 (1.48) | 0.0 (1.0) |
| BNT162b2<br>(for primary vaccine<br>regimens using a<br>subset of data<br>available from<br>February 27, 2021 –<br>September 7, 2021) | 50.3 (17.9) | 0.41 (1.28) | 0.0 (0.0) |
| All mRNA-1273 | 56.6 (18.8) | 0.52 (1.46) | 0.0 (1.0) |
| mRNA-1273<br>(for primary vaccine<br>regimens using a<br>subset of data<br>available from<br>February 27, 2021 –<br>September 7, 2021) | 55.3 (18.3) | 0.48 (1.39) | 0.0 (1.0) |

IQR, interquartile range; SD, standard deviation.

**Table 3.** ORs and 95% CI assessing durability of baseline vaccine protection against infections, hospitalizations, and ICU admissions for each vaccine.

|  | Breakthrough Infection<br>OR* (95% CI) | Hospitalization<br>OR* (95% CI) | ICU Admissions<br>OR*<sup>†</sup> (95% CI) |
| --- | --- | --- | --- |
| **Ad26.COV2.S**<br>(Data available from February 27, 2021<br>– September 7, 2021) |  |  |  |
| Month 1 | 1 (Reference) | 1 (Reference) | 1 (Reference) |
| Month 2 | 1.03 (0.94, 1.14) | 1.01 (0.76, 1.34) | 0.96 (0.39, 2.41) |
| Month 3 | 0.99 (0.89, 1.11) | 1.15 (0.83, 1.60) | 1.89 (0.65, 5.51) |
| Month 4 | 1.16 (1.04, 1.29) | 1.11 (0.79, 1.56) | 1.40 (0.43, 4.55) |
| Month 5 or after | 1.31 (1.18, 1.47) | 1.25 (0.86, 1.80) | .. |
| **BNT162b2**<br>(Data available from February 27, 2021<br>– September 7, 2021) |  |  |  |
| Month 1 | 1 (Reference) | 1 (Reference) | 1 (Reference) |
| Month 2 | 1.21 (1.13, 1.31) | 1.10 (0.88, 1.37) | 0.89 (0.52, 1.51) |
| Month 3 | 1.57 (1.46, 1.70) | 1.45 (1.13, 1.86) | 0.63 (0.31, 1.27) |
| Month 4 | 1.88 (1.74, 2.04) | 1.86 (1.43, 2.43) | 1.62 (0.76, 3.45) |
| Month 5 or after | 2.20 (2.01, 2.40) | 2.38 (1.79, 3.17) | .. |
| **mRNA-1273**<br>(Data available from February 27, 2021<br>– September 7, 2021) |  |  |  |
| Month 1 | 1 (Reference) | 1 (Reference) | 1 (Reference) |
| Month 2 | 1.13 (1.04, 1.23) | 1.03 (0.82, 1.28) | 0.67 (0.39, 1.16) |
| Month 3 | 1.38 (1.26, 1.51) | 1.04 (0.80, 1.34) | 0.54 (0.26, 1.12) |
| Month 4 | 1.77 (1.61, 1.94) | 1.17 (0.89, 1.54) | 1.03 (0.44, 2.41) |
| Month 5 or after | 2.07 (1.87, 2.29) | 1.32 (0.98, 1.79) | .. |
| **BNT162b2** |  |  |  |
| (Data available from January 1, 2021 – September 7, 2021) |  |  |  |
| Month 1 | 1 (Reference) | 1 (Reference) | 1 (Reference) |
| Month 2 | 1.28 (1.21, 1.36) | 1.26 (1.10, 1.44) | 1.20 (0.80, 1.80) |
| Month 3 | 1.68 (1.58, 1.78) | 1.81 (1.55, 2.12) | 0.73 (0.44, 1.21) |
| Month 4 | 1.99 (1.87, 2.12) | 2.45 (2.05, 2.92) | 1.36 (0.80, 2.30)<sup>†</sup> |
| Month 5 | 2.37 (2.21, 2.54) | 3.54 (2.93, 4.27) |  |
| Month 6 or after | 2.93 (2.72, 3.15) | 3.97 (3.26, 4.83) |  |
| **mRNA-1273** |  |  |  |
| (Data available from January 1, 2021 – September 7, 2021) |  |  |  |
| Month 1 | 1 (Reference) | 1 (Reference) | 1 (Reference) |
| Month 2 | 1.21 (1.13, 1.30) | 1.08 (0.91, 1.29) | 0.62 (0.40, 0.97) |
| Month 3 | 1.49 (1.38, 1.61) | 1.23 (1.01, 1.51) | 0.73 (0.43, 1.23) |
| Month 4 | 1.83 (1.68, 1.98) | 1.41 (1.12, 1.77) | 1.17 (0.64, 2.13)<sup>†</sup> |
| Month 5 | 2.15 (1.97, 2.35) | 1.62 (1.27, 2.07) | ... |
| Month 6 or after | 2.76 (2.51, 3.04) | 1.66 (1.26, 2.19) | ... |

CI, confidence interval; ICU, intensive care unit; OR, odds ratio.
\*ORs are matched and conditioned on age group, sex, Gagne comorbidity index score category, three-digit zip, and calendar date.
<sup>†</sup>ICU outcome final time category is month 4 or after.

To estimate the baseline odds of each outcome (separately for each vaccine) we fitted a logistic regression model using all individuals vaccinated with the vaccine of interest who also met all other inclusion and exclusion criteria. We used this fitted logistic regression equation to find the odds of infection among all vaccinated individuals in the first month of follow-up (baseline). To harmonize baseline odds to the unvaccinated population, we adjusted the model for age, sex, comorbidity score, and geographic location (state) according to the unvaccinated population demographics. Specifically, we categorized age (to 18–28, …, 89–98 categories) and comorbidity score (to –2–0, 0–2, 2–5, 5–25 categories) and calculated the proportion of unvaccinated individuals in each age, comorbidity score, sex, and state category. We used these categorical distributions to marginalize the estimated conditional probability from the fitted logistic regression equation to calculate the weighted probability of getting an outcome of interest in the first month, where the weighting was according to the unvaccinated population demographics. Under assumptions of independence the weighted probability of getting an outcome:

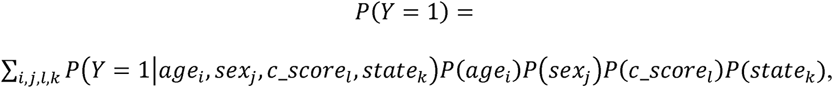

where Y = 1 indicates that the outcome is observed.

Finally, we calculated the baseline odds as the weighted probability of having the outcome of interest in the first month over one minus that probability:

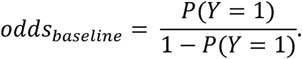

Next, we multiplied the baseline odds of outcome (among all vaccinated with the vaccine of interest) by the OR for the second, third, fourth, fifth, and sixth months of follow-up from the conditional logistic regression models used in the main analysis to approximate the odds of a positive test in the second, third, fourth, fifth, and sixth months respectively:

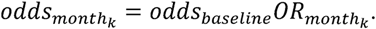

We used the odds of outcome in each month to calculate the probability of that outcome in the one-month period to approximate the incidence proportion for each month for the outcome of interest among those who had received the vaccine of interest:

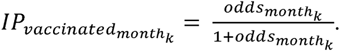

We next estimated the incidence proportion among the unvaccinated. First, we calculated IP for each calendar month in follow-up (February–August). Then, we calculated the proportion of cases in each month of follow-up by the calendar month in the vaccinated cohort (separately for each vaccine) and used these proportions as weights to calculate IP in the unvaccinated for each month of follow-up separately:

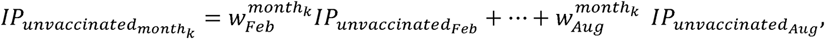

where 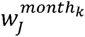 - is the proportion of cases in month *k* of follow-up that happened in calendar month *J*. For example, consider Ad26.COV2.S vaccinated cohort infections outcome. There were 15% of cases in July and 85% in August in the month 5 since follow-up. Then, we calculate the IP for unvaccinated for month 5 of follow-up as

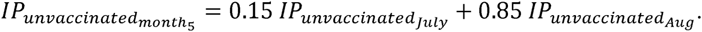

We calculated the VE for each month as one minus the ratio of incidence proportions for the vaccinated as compared to the unvaccinated:

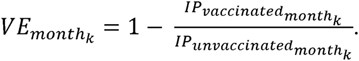

Confidence intervals (95%) for VE estimates were obtained by bootstrapping resampling over 1000 replicates (*41*).

### Sensitivity analyses

To assess durability of protection in groups at relatively higher or lower risk, we stratified cohorts by age and comorbidity score. Due to the low numbers of ICU admission cases, we restricted all sensitivity analyses to infections and hospitalizations. For age-stratified analyses, we repeated the original analysis using the same procedures and methods and built models separately for study participants 65 years and older and younger than 65 years of age. Similarly, for comorbidity-stratified analyses, we repeated our analyses for study participants with a comorbidity score of less than two and a comorbidity score of two or greater.

## Results

### Vaccine cohorts, cases, and controls

Individuals fully vaccinated with one of the three authorized or approved vaccines in the United States and meeting study inclusion and exclusion criteria represented the broader vaccinated cohorts from which our matched case-control samples were drawn (see attrition flowchart, Fig. S1). Together, 17,017,435 individuals were included in these cohorts, representing close to 10% of all individuals fully vaccinated in the United States by the end of our study period, September 7, 2021 (*14*). Overall vaccine cohorts differed in some demographic factors across vaccines (Tables 1 and 2), reflecting differences in vaccine authorization and eligibility timing, as well as differences in distribution and availability. Vaccine cohorts where the last dose was on or after February 27, 2021 were more similar. Among those, mean age was older in mRNA-1273 (55.3 years) relative to Ad26.COV2.S (50.9 years) and BNT162b2 (50.3 years). More individuals had high (≥ 2) comorbidity scores in mRNA-1273 (12.4%) than BNT162b2 (10.6%) and Ad26.COV2.S (10.5%), as well. Ad26.COV2.S had fewer women and more individuals insured through Medicaid (52% and 8.7%, respectively) than BNT162b2 (57% and 7.2%) or mRNA-1273 (57% and 7.3%). The number of cases and matched controls for each outcome for each vaccine are listed in Table S1. Across vaccines and outcomes, the included cases represented 98% of all eligible cases. Compared to those vaccinated on or after February 27, cases among those who were vaccinated earlier (from January 1 to February 27) had substantially more comorbidities (Gagne score ≥ 4, 26.1% vs 7.3%) and were older (age ≥ 85, 28.7% vs 3.2%) (Table S2).

### Change in effectiveness of Ad26.COV2.S over time

Among those who were vaccinated with the Ad26.COV2.S vaccine, modest waning of protection against infection was observed by month 5+ as measured by the odds ratios (OR [95% CI], 1.31 [1.18–1.47]; Table 3 and Fig. 1A). Corresponding calculated VE was stable across the study period. Calculated VE was 74% (95% CI, 72–75) at month 1 and 74% (95% CI, 70–76) at month 5 or any time after (Table 4 and Fig. 2A). No evidence of waning protection against hospitalization was observed (OR for month 5+ = 1.25 [95% CI, 0.86–1.80]). Translated VE ranged from 81% (95% CI, 76–82) at month 1 to 77% (95% CI, 64–83) at month 5 or after. For ICU admission, protection did not wane over time. Similarly, when stratifying by age or comorbidity score, stable protection against infection and hospitalization was observed across strata (Figs. S2–S5).

**Fig. 1A.**
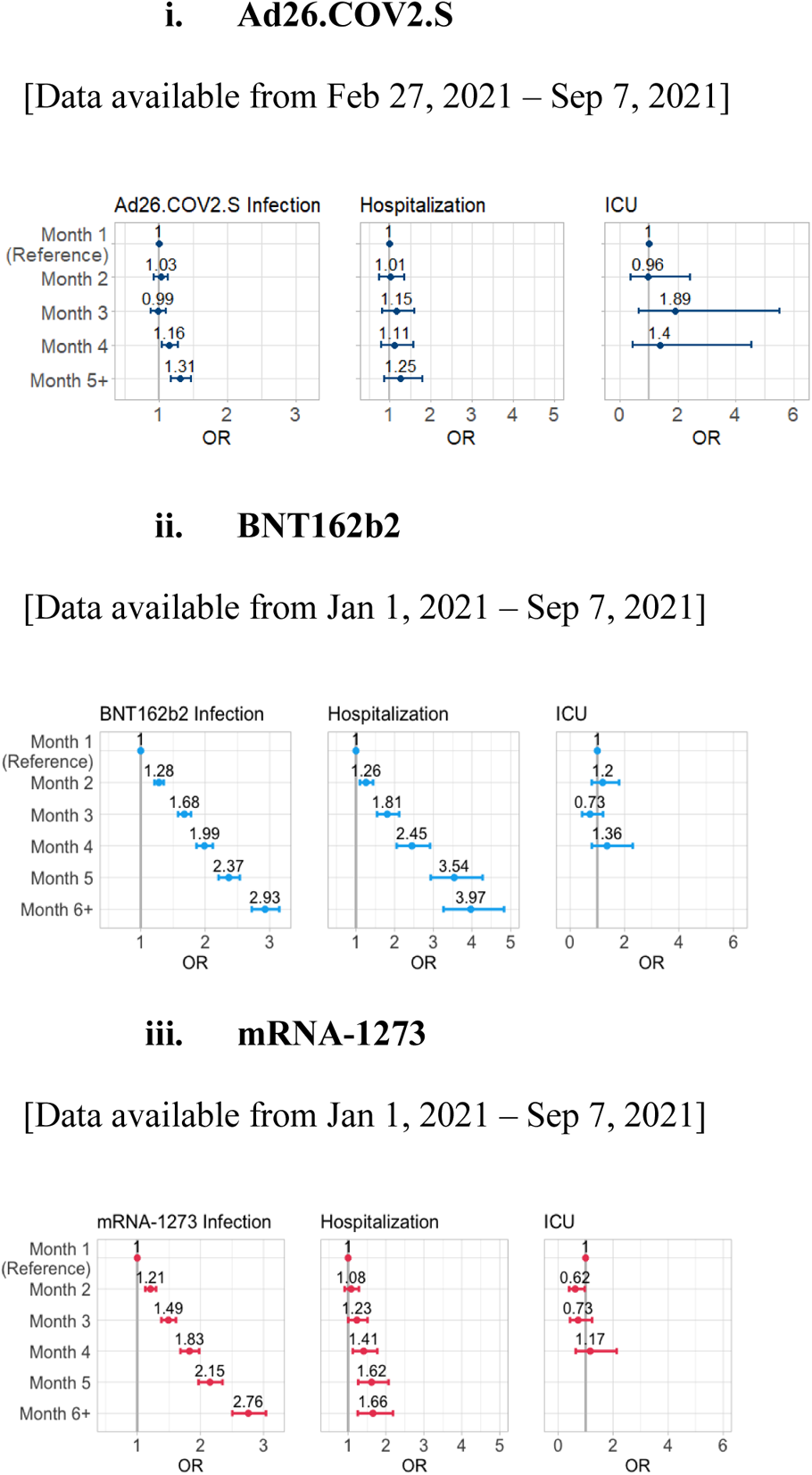
OR and 95% CI assessing durability of baseline vaccine protection against infections, hospitalizations, and ICU admissions leveraging all available data for each vaccine.

**Fig. 2A.**
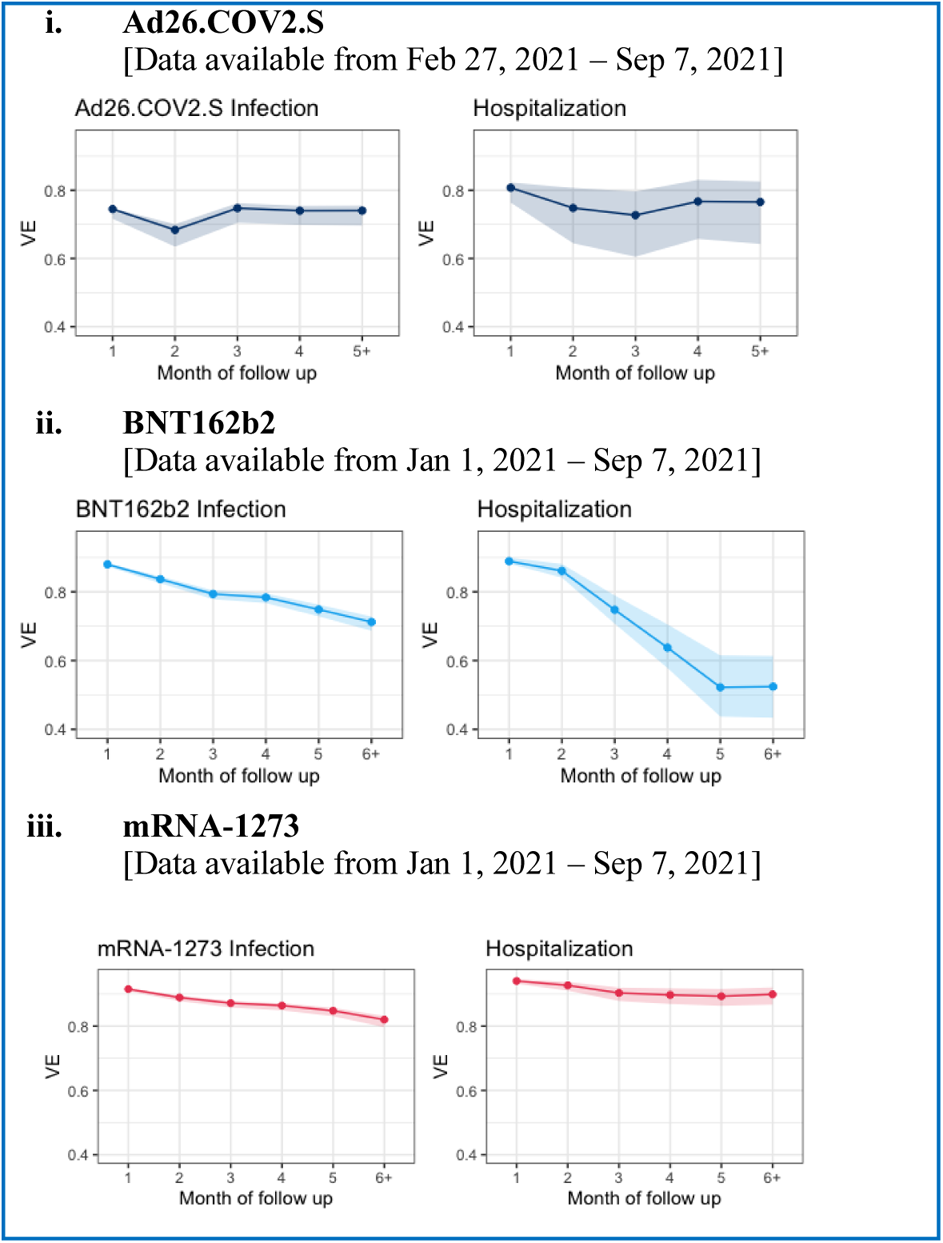
Estimated VE and 95% CI against infections and hospitalizations by month of follow-up for primary vaccine regimens using all available data.

**Table 4.** Estimated VE against infections and hospitalizations by month of follow-up.

|  | **Breakthrough Infection**<br>**VE (95% CI)** | **Hospitalization**<br>**VE (95% CI)** |
| --- | --- | --- |
| **Ad26.COV2.S**<br>(Data available from February 27, 2021 –<br>September 7, 2021) |  |  |
| Month 1 | 0.74 (0.72, 0.75) | 0.81 (0.76, 0.82) |
| Month 2 | 0.68 (0.63, 0.70) | 0.75 (0.64, 0.81) |
| Month 3 | 0.75 (0.70, 0.76) | 0.73 (0.61, 0.80) |
| Month 4 | 0.74 (0.70, 0.75) | 0.77 (0.66, 0.83) |
| Month 5 or after | 0.74 (0.70, 0.76) | 0.77 (0.64, 0.83) |
| **BNT162b2**<br>(Data available from February 27, 2021 –<br>September 7, 2021) |  |  |
| Month 1 | 0.89 (0.88, 0.89) | 0.92 (0.91, 0.93) |
| Month 2 | 0.86 (0.84, 0.87) | 0.90 (0.87, 0.92) |
| Month 3 | 0.86 (0.84, 0.87) | 0.87 (0.84, 0.90) |
| Month 4 | 0.86 (0.84, 0.86) | 0.86 (0.82, 0.89) |
| Month 5 or after | 0.84 (0.82, 0.85) | 0.84 (0.79, 0.88) |
| **mRNA-1273**<br>(Data available from February 27, 2021 –<br>September 7, 2021) |  |  |
| Month 1 | 0.92 (0.91, 0.92) | 0.94 (0.93, 0.95) |
| Month 2 | 0.90 (0.89, 0.90) | 0.93 (0.91, 0.94) |
| Month 3 | 0.90 (0.89, 0.91) | 0.93 (0.90, 0.94) |
| Month 4 | 0.89 (0.88, 0.90) | 0.93 (0.91, 0.95) |
| Month 5 or after | 0.88 (0.87, 0.89) | 0.93 (0.91, 0.95) |
| **BNT162b2**<br>(Data available from January 1, 2021 –<br>September 7, 2021) |  |  |
| Month 1 | 0.88 (0.87, 0.88) | 0.89 (0.88, 0.90) |
| Month 2 | 0.84 (0.83, 0.84) | 0.86 (0.84, 0.88) |
| Month 3 | 0.79 (0.78, 0.80) | 0.75 (0.71, 0.79) |
| Month 4 | 0.78 (0.77, 0.80) | 0.64 (0.58, 0.71) |
| Month 5 | 0.75 (0.73, 0.76) | 0.52 (0.44, 0.61) |
| Month 6 or after | 0.71 (0.69, 0.73) | 0.52 (0.43, 0.61) |
| **mRNA-1273**<br>(Data available from January 1, 2021 –<br>September 7, 2021) |  |  |
| Month 1 | 0.92 (0.91, 0.92) | 0.94 (0.93, 0.95) |
| Month 2 | 0.89 (0.88, 0.90) | 0.93 (0.91, 0.94) |
| Month 3 | 0.87 (0.86, 0.88) | 0.90 (0.88, 0.92) |
| Month 4 | 0.86 (0.85, 0.87) | 0.90 (0.87, 0.92) |
| Month 5 | 0.85 (0.83, 0.86) | 0.89 (0.86, 0.92) |
| Month 6 or after | 0.82 (0.80, 0.83) | 0.90 (0.87, 0.92) |

CI, confidence interval; VE, vaccine effectiveness.

### Change in effectiveness of BNT162b2 over time

Among all those vaccinated with BNT162b2 (dose 1 ≥ January 1, 2021), the odds of a breakthrough infection were successively higher for each month of follow-up (OR for month 6+ of BNT162b2 = 2.93 [95% CI, 2.72–3.15]; Table 3 and Fig. 1A) and translated VE ranged from 88% (95% CI, 87–88) at month 1 to 71% (95% CI, 69–73) at month 6 or after (Table 4 and Fig. 2A). Odds of hospitalization increased considerably over time (OR for month 6+ = 3.97 [95% CI, 3.26–4.83]; Table 3 and Fig. 1A) and VE against hospitalization ranged from 89% (95% CI, 88–90) at month 1 to 52% (95% CI, 43–61) at month 6 or after (Table 4 and Fig. 2A). There was no evidence of waning protection against ICU admissions at any point.

Among a subset who were vaccinated with BNT162b2 (dose 2 on or after February 27, 2021), the odds of a breakthrough infection were successively higher for each month of follow-up (OR for month 5+ of BNT162b2 = 2.20 [95% CI 2.01–2.40]; Table 3 and Fig. 1B) and corresponding translated VE ranged from 89% (95% CI, 88–89) at month 1 to 84% (95% CI, 82–85); Table 4 and Fig. 2B). For hospitalizations, odds increased successively (OR for month 5+ = 2.38 [95% CI, 1.79–3.17]; Table 3 and Fig. 1B) and translated VE ranged from 92% (95% CI, 91–93) at month 1 to 84% (95% CI, 79–88) at month 5 or after (Table 4 and Fig. 2B). In age- and comorbidity-stratified groups, protection against infection and hospitalization protection waned similarly in all groups and was consistent with the effects observed in the overall BNT162b2 cohort (Figs. S2–S5).

**Fig. 1B.**
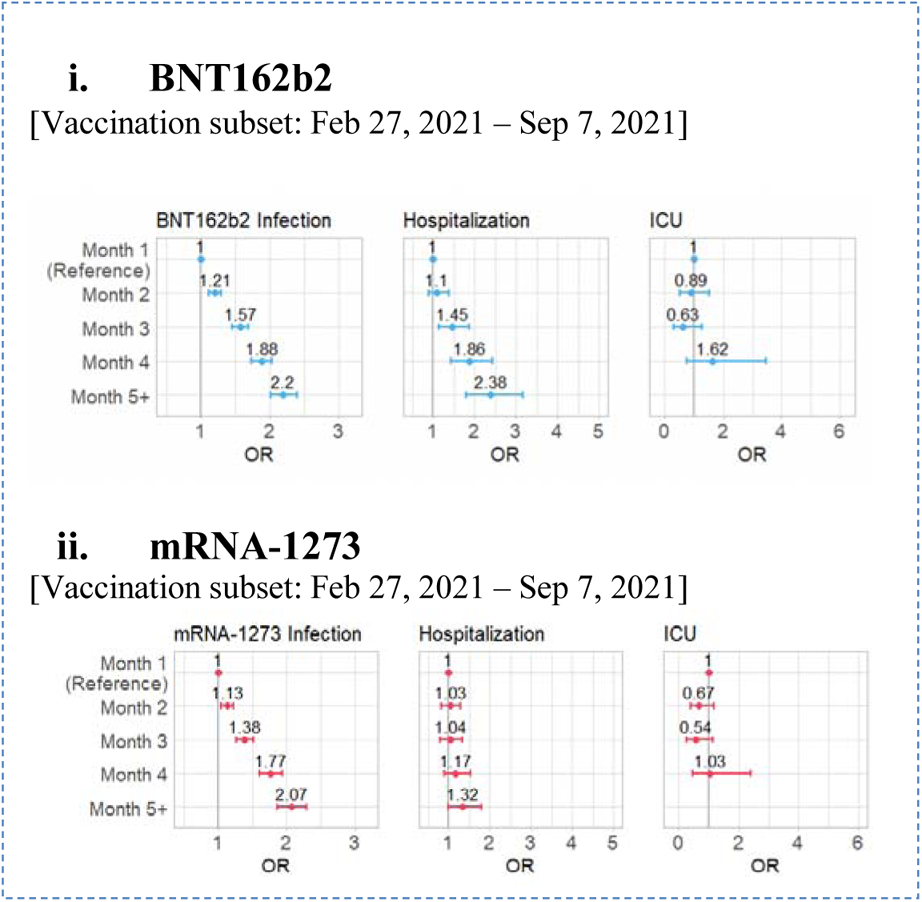
ORs and 95% CI assessing durability of baseline vaccine protection against infections, hospitalizations, and ICU admissions using a subset of data available from February 27, 2021 – September 7, 2021.

**Fig. 2B.**
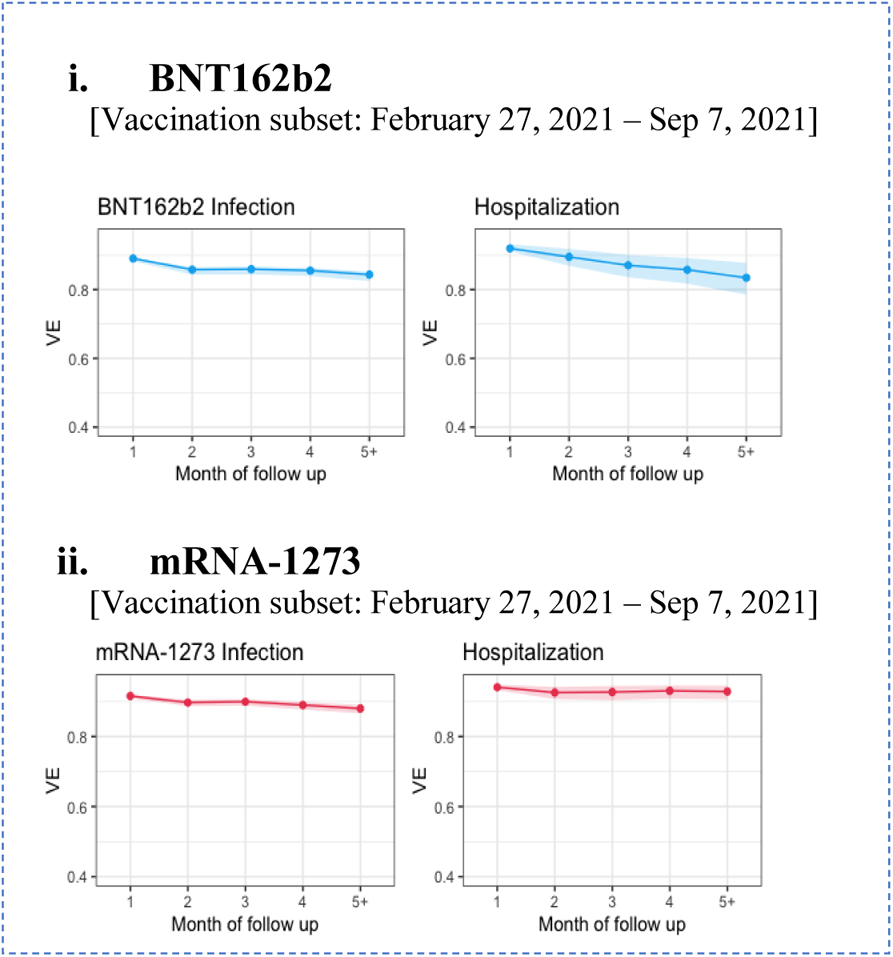
Estimated VE and 95% CI against infections and hospitalizations by month of follow-up for primary vaccine regimens using a subset of data available from Februar*y* 27, 2021 – September 7, 2021.

### Change in effectiveness of mRNA-1273 over time

Among all those vaccinated with mRNA-1273 (dose 1 ≥ January 1, 2021), odds of a breakthrough infection were successively higher for each month of follow-up (OR for month 6+ of mRNA-1273 = 2.76 [95% CI, 2.51–3.04]; Table 3 and Fig. 1A) and corresponding translated VE against infection ranged from 92% (95% CI, 91–92) at month 1 to 82% (95% CI, 80–83) (Table 4 and Fig. 2A). Odds of hospitalization increased modestly (OR for month 6+ = 1.66, 95% CI [1.26–2.19]) and translated VE ranged from 94% (95% CI, 93–95) at month 1 to 90% (95% CI, 87–92). There was no evidence of waning protection against COVID-19-related ICU admissions at any point.

Among a subset who were vaccinated with the mRNA-1273 vaccines (dose 2 ≥ February 27, 2021), the odds of a breakthrough infection were successively higher for each month of follow-up (OR for month 5+ of mRNA-1273 = 2.07 [95% CI, 1.87–2.29]; Table 3 and Fig. 1B). Corresponding VE against infection ranged from 92% (95% CI, 91–92) at month 1 to 88% (95% CI, 87–89) at month 5 or after (Table 4 and Fig. 1B). Protection against hospitalization did not wane over time (OR for month 5+ = 1.32 [95% CI, 0.98–1.79]) and translated VE was stable ranging from 94% (95% CI, 93–95) at month 1 to 93% (95% CI, 91–95).

In age- and comorbidity-stratified groups, protection against infection and hospitalizations, modest waning was observed in higher-risk groups, but not in lower-risk groups (Figs. S2–S7).

## Discussion

Using claims data for over 168 million individuals in the United States from January 1 to September 7, 2021, we used a matched case-control study design to assess the durability of protection against breakthrough infections, hospitalizations, and ICU admissions over time from full vaccination. The database used in this study was representative of the proportions of payer type used by the broader US population (*14, 15*). Although unable to distinguish between declining effectiveness due to immunological processes or the emergence of novel variants, the study provides insights into real-world outcomes of vaccines over time. By matching cases and controls for each vaccine on confounding variables including calendar time and location, any impact of viral strain and the variable background rates of infection would likely be accounted for (*16*). The study was designed to compare each vaccine to itself and therefore direct comparison between vaccines should be avoided, as no formal matching or adjustment for baseline differences was made.

Although, as seen in randomized trials, the level of efficacy of a single-dose Ad26.COV2.S vaccine was lower than that seen with two-dose mRNA vaccines, we found that the level of protection after vaccination with the Ad26.COV2.S vaccine was durable for both breakthrough infections and hospitalizations. For both mRNA vaccines, protection against infections waned in all considered cohorts and sub-groups (age- and comorbidity-stratified). Protection against hospitalization was generally stable for mRNA-1273 in younger, healthier cohorts, and waned modestly in higher-risk groups. For BNT162b2, protection against hospitalization waned in all cohorts and sub-groups. All vaccines showed durable protection over the study period against ICU admission, showing sustained protection against critically severe disease.

Recent studies have suggested a more stable neutralizing antibody response for those inoculated with the Ad26.COV2.S vaccine as compared to the BNT162b2 and mRNA-1273 vaccines, potentially due to differences in the mechanism of action (*17, 18*), which may translate into more durable protection. Results from this study corroborate this deduction. However, any statement about the durability of vaccine effectiveness should be interpreted with caution due to potential floor or ceiling effects where a more effective vaccine has more room for waning compared to a less effective one.

A strength of our study design comparing the odds of outcome events from later periods to a period soon after full vaccination is that it avoids the need for an unvaccinated control group, which in national insurance claims databases may be subject to confounding and underreporting (*4*). However, as the observed ORs can be difficult to interpret, we incorporated assumptions about the incidence rate in the unvaccinated population during the study period, and indirectly estimated baseline and subsequent VE for each vaccine and outcome (Materials and Methods and Supplementary Text). Our translated baseline VE estimates were consistent with the published literature for the three vaccines primary series showing mRNA-1273 to have the highest level of protection, followed by BNT162b2, and by single-dose Ad26.COV2.S, adding face validity to the method and assumptions of the VE translation (*1–8, 19*).

Studies specifically examining durability of vaccine effectiveness have been limited. A recent metanalysis of studies evaluating durability defined as change over time in VE identified 78 studies (BNT162b2, n=38; mRNA-1273, n=23; Ad26.COV2.S, n=9; and AstraZeneca-Vaxzevria, n=8) (*20*). The study concluded that, on average for all four vaccines, vaccine efficacy or effectiveness for symptomatic COVID-19 decreased by 24.9% (95% CI, 13.4–41.6) in people of all ages. For severe COVID-19, vaccine efficacy or effectiveness decreased by 10.0% (95% CI, 6.1–15.4) in people of all ages. In this metanalysis, the studies of Ad26.COV2.S durability of protection were limited in number and sample size and hence inconsistent especially with regards to durability of protection against breakthrough infections. For example, a study by Lin and colleagues (*21*) found that for breakthrough infections, VE declined from 74.8% (95% CI, 72.5–76.9) at month 1 to 64.0% (95% CI, 60.3–67.4) at month 6. For hospitalization, VE remained stable, ranging from 77.7% (95% CI, 68.0–84.5) in month 1, to 81.7% (95% CI, 68.6–89.3) in month 6. A study by Rosenberg showed no time trend or waning of effectiveness for hospitalizations (RR 0.82 [95% CI, 0.63–1.07]) (*9*). More recently, data from the CDC on COVID-19 cases by vaccine product shows those receiving single-dose Ad26.COV2.S to have lower incidence proportions of cases starting in January 2022 (*22*). However, it is not clear if this unadjusted data is driven by differential protection against the Omicron variant or differential waning due to immunological processes. On the other hand, one study showed a large decline in protection against breakthrough infections for Ad26.COV2.S ranging from 64% (95% CI, 58–69) on day 14 to 36% (95% CI, 31–42) on day 172 (*23*). Another showed a decline ranging from 52% (95% CI, 44–59) at day <90 from vaccination to 28% (95% CI, –8 to 53) (*24*), however, the confidence interval was very wide due to the limited number of events. The present study confirms the durability of protection of single-dose Ad26.COV2.S against hospitalizations and severe disease.

Results from the present study are consistent with the findings from the metanalysis for BNT162b2 and mRNA-1273 for breakthrough infections and for mRNA-1273 hospitalizations. For BNT162b2 hospitalization, the pattern of declining effectiveness for BNT162b2 from February 27 to the end of the study follow-up is consistent with the published literature. However, this study is the first to report such a large decline in effectiveness leveraging the full study period of January to September. This finding requires further investigation, especially since stratifications by age and comorbidities showed no discernible pattern to the decline in effectiveness. Moreover, the translated VE against hospitalization for BNT162b2 (January to September data) was lower than those observed in clinical trials and other real-world effectiveness studies, which may suggest a potential unobservable difference in this population. The characteristics of the populations of those vaccinated from January 1 to February 27 had substantially more comorbidities (Gagne score ≥6) and were older than those vaccinated from February 27 to September 7. For example, many of the early recipients of BNT162b2 were health care workers who may have been at high risk for viral exposure and transmission due to their occupation. This hypothesis, however, is not testable in insurance claims databases because occupational status is not recorded.

Four large representative studies examined the durability of effectiveness in preventing hospitalization in BNT162b2. One study showed a decline in VE from 91% (95% CI, 88–93) on day 14-120 to 77% (95% CI, 67–84) on day 121 or after (*7*). A study by Tartof and colleagues showed no decline in VE against hospitalization (*5*). A study from Qatar found a decline in VE against hospitalization from 96% (95% CI, 93.9–97.4) at month 1 to 55.6% (95% CI, −44.3 to 86.3) at month 7 or after (*25*). This declining VE was based on very few events (6 among cases and 11 among controls) (*25*) and therefore has very wide confidence intervals. VE against hospitalization remained high at month 6 (VE 88.9% [95% CI, 52.1–97.4]). Moreover, the study by Lin and colleagues found the VE for BNT162b2 against breakthrough infections to decline from 85.5% (95% CI, 85.0–86.0) at month 1 to 67.8% (95% CI, 65.9–69.7) at month 8 (*21*). The VE against hospitalization remained high and there was no discernible pattern of decline. Further population-based studies are needed to understand the drivers of this observed discrepancy in the declining VE for BNT162b2 and to evaluate the long-term durability of protection of the primary series.

Several additional limitations should be considered when interpreting our results. First, although we reduced differences between cases and controls that could affect the probability of COVID-19-related outcomes through matching, there could be remaining unmeasured effects such as occupation-related exposure. The analysis strategy maximized the number of cases by disallowing cases from serving as controls for other cases. This could have induced bias since the experience of the controls is not representative of the true experience of vaccinated individuals. However, simulations of this strategy suggest this bias is limited when event rates are low, as they were in this study. Individuals were included in this database for COVID-19-related activities; as such, the unvaccinated population was restricted to those interacting with the healthcare system for COVID-19-related concerns and thus is not representative of the general population and may have inflated VE estimates. Finally, this study relied primarily on open claims, which means other COVID-19-related medical encounters may have occurred within our population that we did not observe. However, individuals included in our study had observable vaccination records as well as healthcare utilization prior to vaccination, so observability should not be a significant limitation. Finally, this study is restricted in duration to an era before the dominance of the Omicron variant. However, the study remains relevant for understanding the natural course of VE in 2021 and will have implications for 2022 and beyond.

In summary, while the starting protection level provided by the primary series may differ by vaccine type and mechanism of action, this study demonstrated by comparing each vaccine to its own baseline protection that the three vaccines may have unique durability profiles. As the COVID-19 pandemic continues, as low to middle income countries remain largely unvaccinated and as more high income countries implement a standard of care consisting of one or more boosters, further investigation is critical to understand the level of protection and the durability of response over longer periods, novel variants, and in response to homologous and heterologous boosting.

## Data Availability

Upon reasonable request, researchers may get access to the data and analytics infrastructure for prespecified collaborative analyses.

## Acknowledgements

The authors thank the following team members for critical feedback during the development of the study protocol and/or review of findings: Penny Heaton (Janssen), Johan Van Hoof (Janssen), Deb Ricci (Janssen), Ray Harvey (Janssen), Sid Jain (Janssen), Xiaoying Wu (Janssen), Sophia Gallucci (Aetion), Ann Madsen (Aetion), and Mike Batech (Aetion). This work was funded by Janssen Pharmaceuticals Research & Development. Editorial support was provided by Kurt Kunz, MD, MPH, and Jill E. Kolesar, PhD (Cello Health Communications/MedErgy) and funded by Janssen Global Services.

## Author Contributions

Conceptualization: AZ, MO, BN, TK, NK, SS, KS

Data curation: RS, JH

Formal analysis: AZ, MO, NZ

Investigation: AZ, MO

Methodology: AZ, MO, NZ, JH, BN, SS, KS

Validation: AZ, MO, RS, NZ, JH

Visualization: AZ, MO, RS, NZ, MR

Writing – original draft: AZ, MO, RS, NZ, MR, BN, KS

Writing – review & editing: AZ, MO, BN, TK, NK, SS, KS

All authors had full access to all the data in the study and accept responsibility to submit for publication.

## Data and Materials Availability

The de-identified row level data may be obtained through a HealthVerity data license and are not publicly available.

## Supplementary Materials

Supplementary Text

Figs. S1 to S6

Tables S1 to S2

## Supplementary Materials

### Published COVID-19 vaccine durability studies used for comparison to the current study

We compared our estimates of VE to similar studies from the literature and plotted these values for each vaccine and outcome (Figs. S6, S7). For inclusion, prior studies had to meet the following criteria:

(iv) Must be in a published or preprint study
(v) Must report VE on at least one of the three COVID-19 vaccines currently in the United States (Ad26.COV2.S, BNT162b2, and mRNA-1273)
(vi) VE could not be reported in aggregate across vaccines (e.g., VE could not be for “mRNA vaccines,” or “any vaccine” and had to specify a single vaccine)
(viii) VE must come from completed vaccine course (e.g., no VE percentages for initial dose in a two-dose series or booster shots only)
(viii) VE had to have at least two timepoints to qualify as a measure of durability over time
(ix) VE must be for infection or hospitalization outcomes
(x) VE percentage at a specific timepoint had to be listed in a table
(xi) To reduce bias in comparing distinctly different cohorts between our study and others, we required that VE could not come from a special-interest population (e.g., high-risk groups like health care workers or immunocompromised patients) or a particular demographic category (≥65 years of age)

### Supplementary Text

#### Vaccine effectiveness

VE for infection and hospitalizations remained strong for all three vaccines throughout the follow-up period (Table 4). However, these results differed for BNT162b2, and mRNA-1273 when using the full available data (January 1, 2021, to September 7, 2021) or when aligning the cohort entry time to match the entry time of Ad26.COV2.S (February 27, 2021) especially for hospitalization. Leveraging the full available data, for infections, VE in the first month of follow-up was 0.74, 0.88, and 0.92 for Ad26.COV2.S, BNT162b2, and mRNA-1273 vaccines, respectively. Ad26.COV2.S showed stable VE over time. The mRNA vaccines had decreased VE by the final month of follow-up with BNT162b2 decreasing to 0.71 and mRNA-1273 decreasing to 0.82 in month 6+. Similar trends were observed when aligning the cohort entry date to February 27, 2021.

For COVID-19-related hospitalizations, using all available data, VE in the first month of follow-up was 0.81, 0.89, and 0.94 for Ad26.COV2.S, BNT162b2, and mRNA-1273 vaccines, respectively. In month 5+, Ad26.COV2.S VE against hospitalization was 0.76, and for BNT162b2, and mRNA-1273 in the 6+ month of follow-up VE against hospitalization was 0.52 and 0.90 respectively. When aligning the cohort entry date to February 27, 2021, for BNT162b2, and mRNA-1273, the waning in VE for BNT162b2 became less pronounced. Trends for mRNA-1273 VE remained similar to those previously observed.

These results confirm and extend estimations of peak VE (i.e., in the first month after full vaccination) for all three vaccines separately against infections and severe disease although the calculated VE against hospitalization for BNT162b2 was lower than the VE reported in clinical trials and other real-world effectiveness studies. The observed waning in VE against hospitalization for BNT162b2 requires further discussion. The apparent clear decline in VE over time is observed when the study cohort includes subjects vaccinated in January and February 2021. This may be driven, at least in part, by waning immune responses in older and more comorbid patients (Tables 1, 2, S2).

Moreover, subgroups may experience differential waning. For example, vaccine administration was initially prioritized for health care workers and residents of long-term care facilities and these individuals might account for the differences in durability estimates observed when those vaccinated in January and February 2021 were included in the BNT162b2 cohort. These individuals may have had great SARS-CoV-2 exposure, more frequent testing, or different immune responses (in the case of long-term care residents) as compared to other individuals. Further investigation is needed to fully understand the complete durability profile of each of the vaccines.

Our study population is consistent with previously published trends in one of the largest samples to date, including approximately 10% of the fully vaccinated individuals in the United States during our study period (*14*). This is of particular importance for the evaluation of the Ad26.COV2.S vaccine, which has generally been assessed with much smaller samples relative to the mRNA vaccines or not at all. Our methods for translating OR to VE come with several limitations. First, we estimated incidence proportion from large cohorts of vaccinated and unvaccinated individuals where absolute case numbers likely represent underestimates, in part due to the potential for asymptomatic and unconfirmed infections (*42–44*). While this is not an issue if this is equally true for both vaccinated and unvaccinated groups, if providers tested or hospitalized individuals differently based on vaccine status, this may result in bias. Second, many COVID-19-related encounters in the United States, including vaccination and tests, occurred in settings where they were not billed to any payer (e.g., mass vaccination clinics) and thus cannot be observed in claims. This can affect the results primarily via misclassification of unvaccinated individuals. This issue would be expected to result in an underestimate of VE (*4, 45*). Finally, the cohorts differ by the timing of events, and relatedly, their demographic makeup. As described above, we attempted to harmonize the baseline odds according to the demographic composition of the unvaccinated cohort. However, demographic differences persist and may limit the comparability of VE across vaccines.

---

**Fig. S1.**
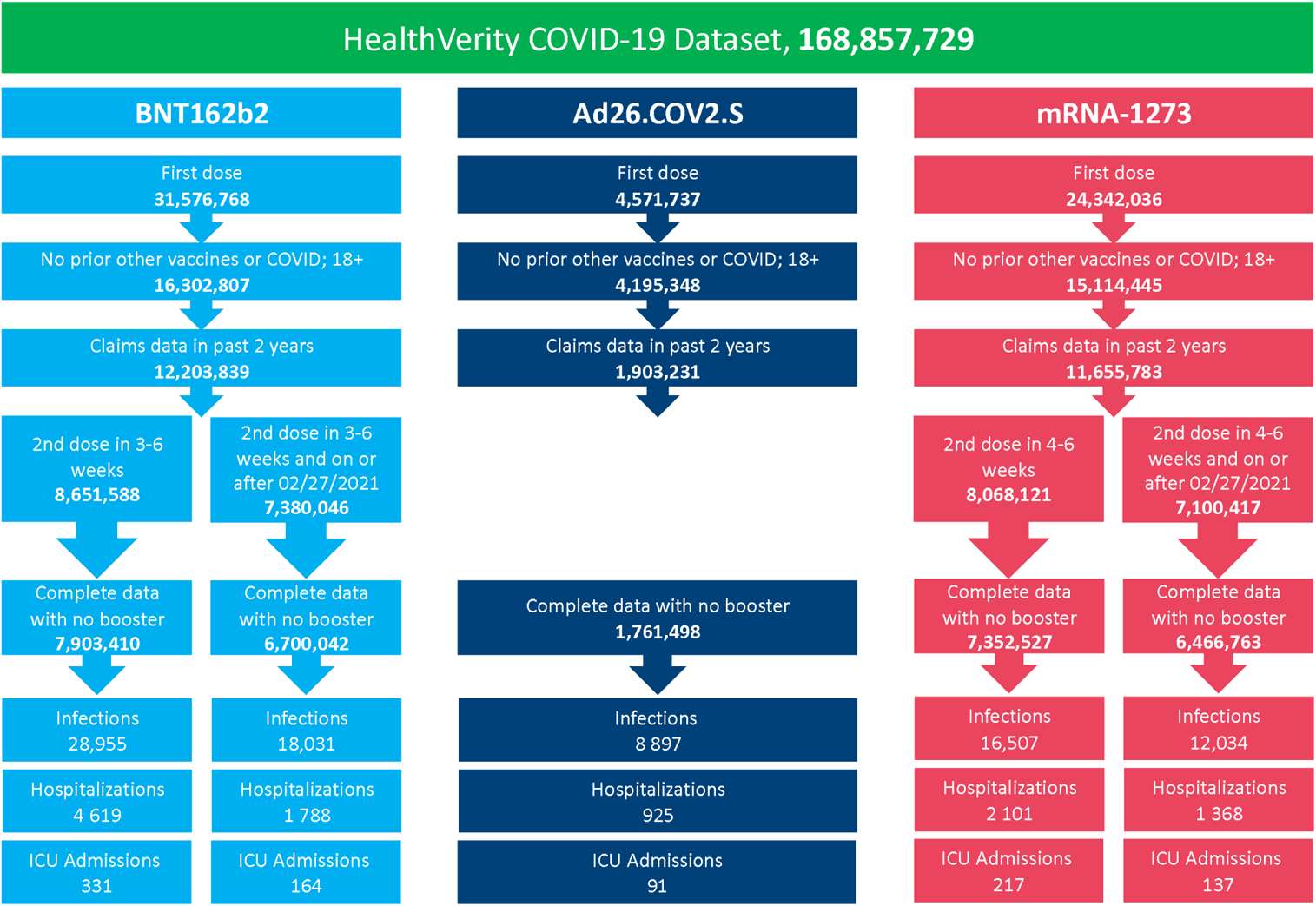
Study inclusion flowchart. ICU=intensive care unit.

**Fig. S2.**
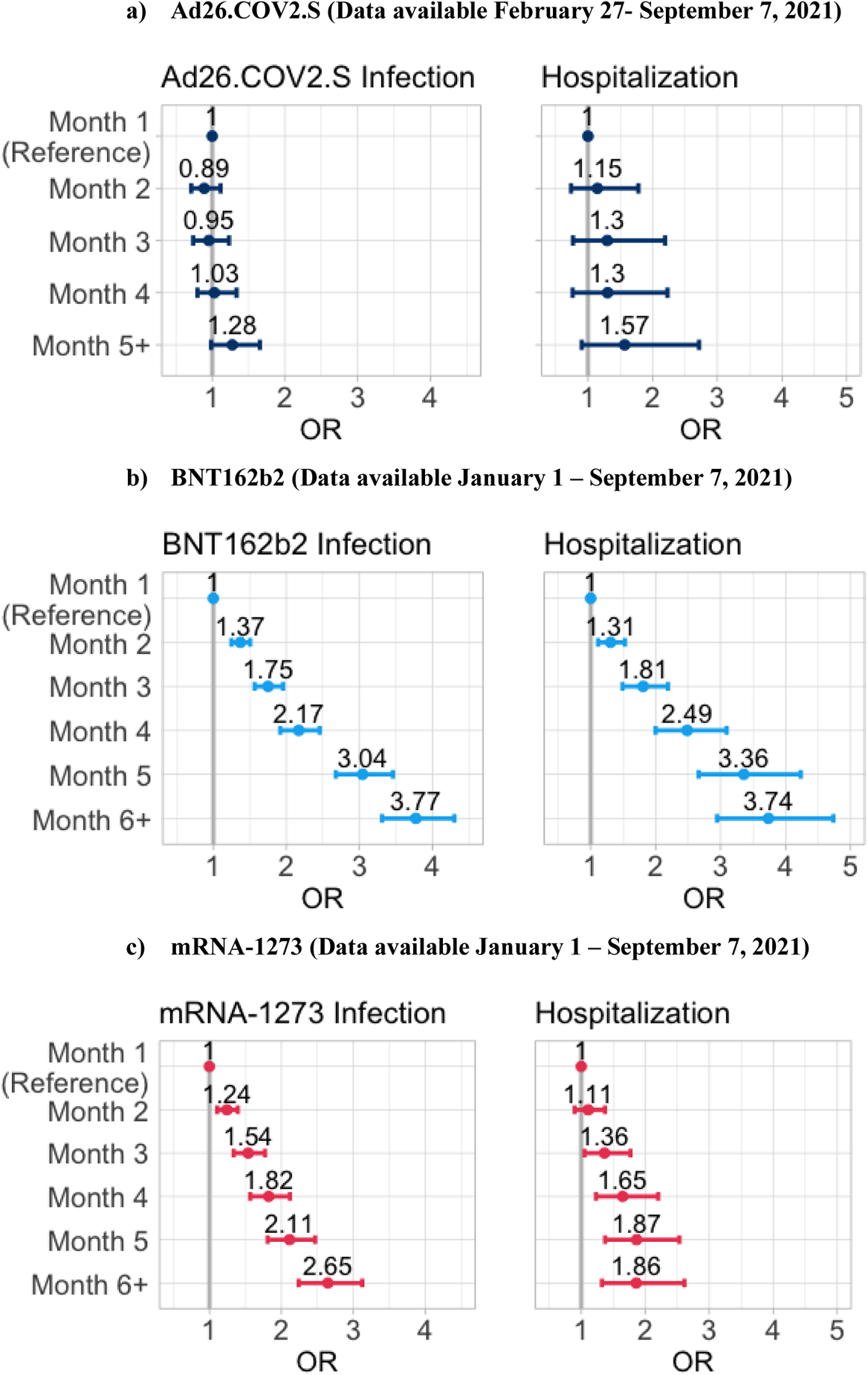
OR and 95% CI assessing durability of baseline vaccine protection against infections and hospitalizations in persons age 65 years or older. CI=confidence interval. OR=odds ratio.

**Fig. S3.**
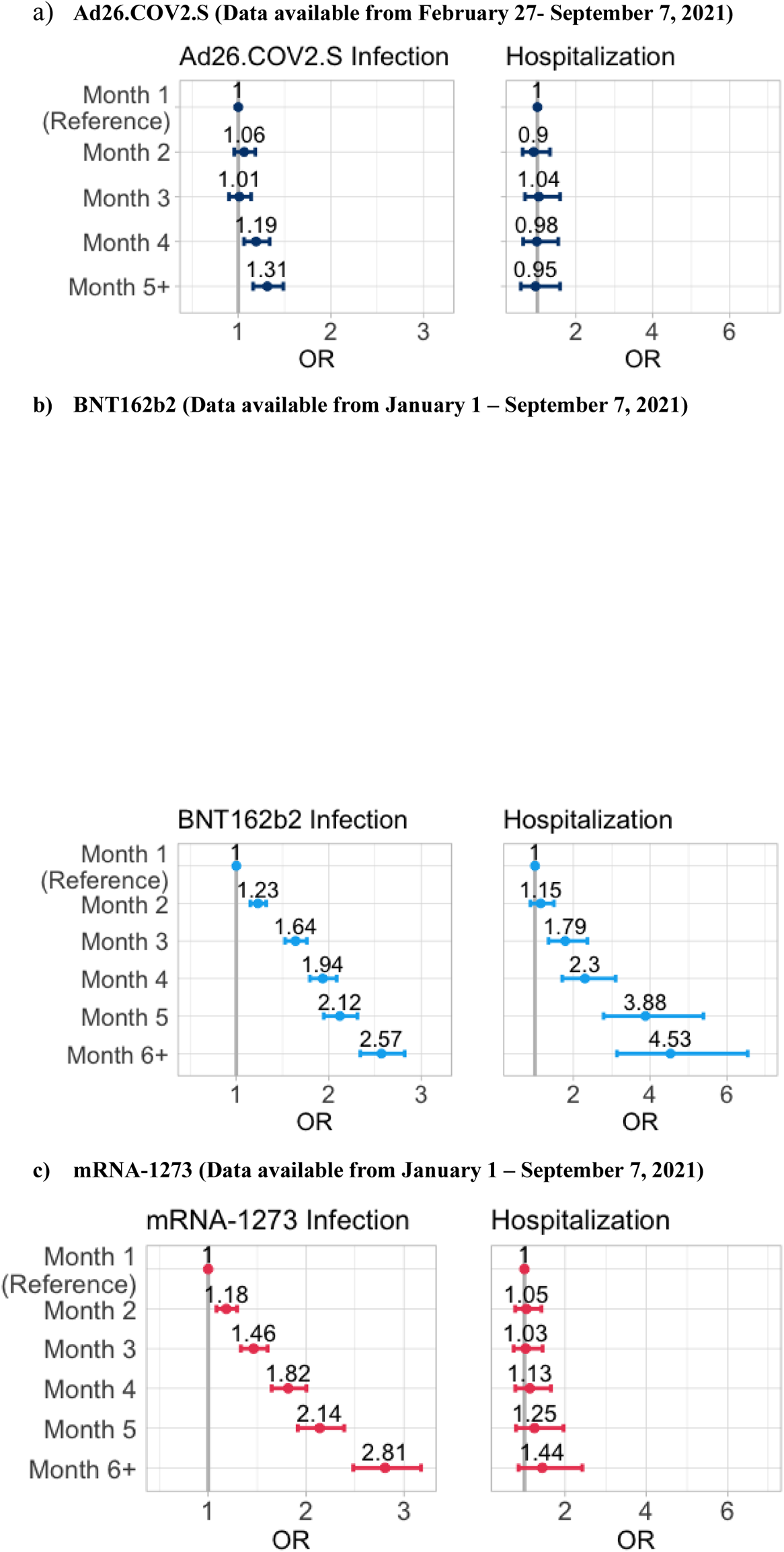
OR and 95% CI assessing durability of baseline vaccine protection against infections and hospitalizations in persons younger than 65 years of age. CI=confidence interval. OR=odds ratio.

**Fig. S4.**
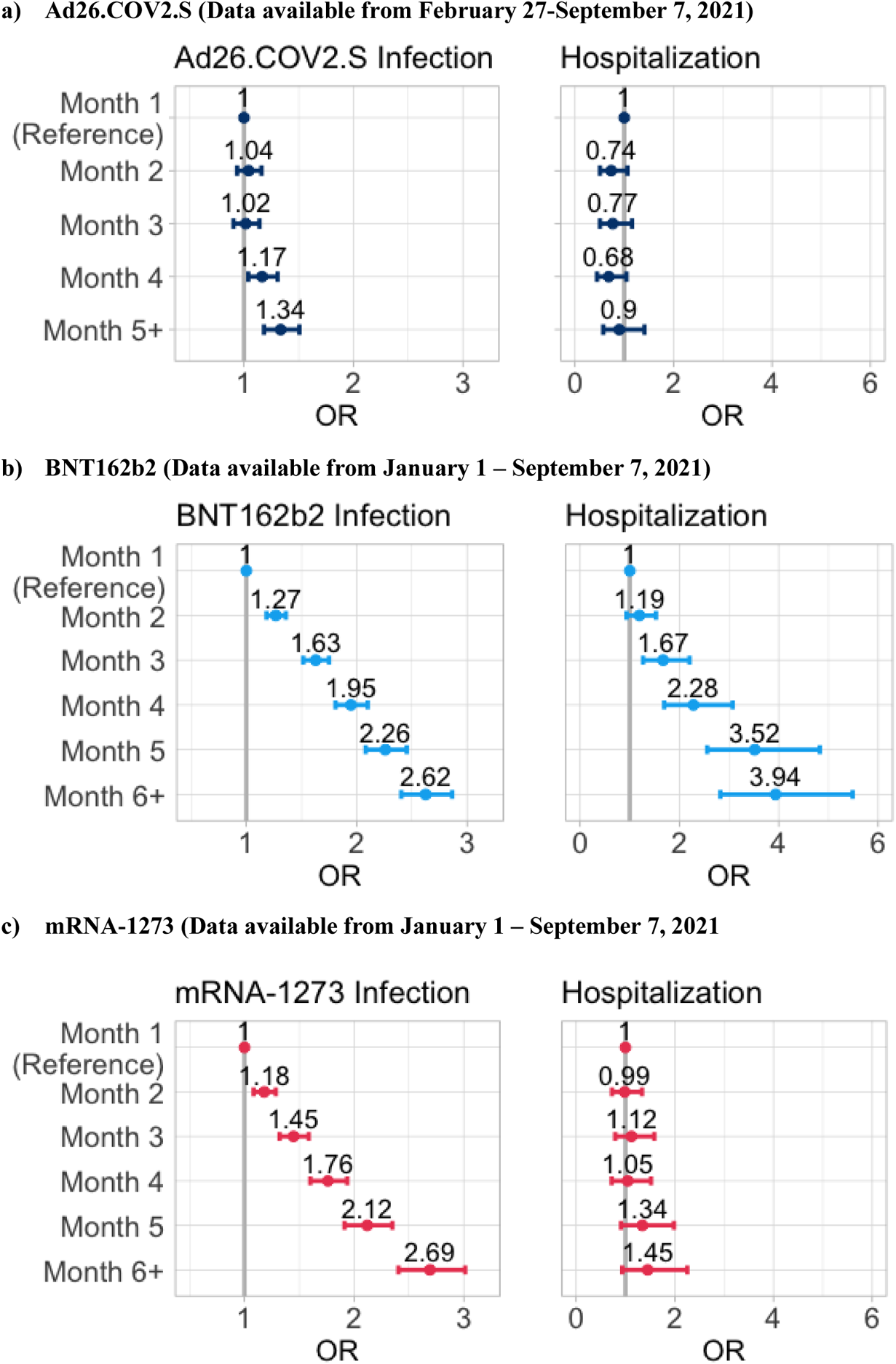
OR and 95% CI assessing durability of baseline vaccine protection against infections and hospitalizations in persons with comorbidity score less than 2. CI=confidence interval. OR=odds ratio.

**Fig. S5.**
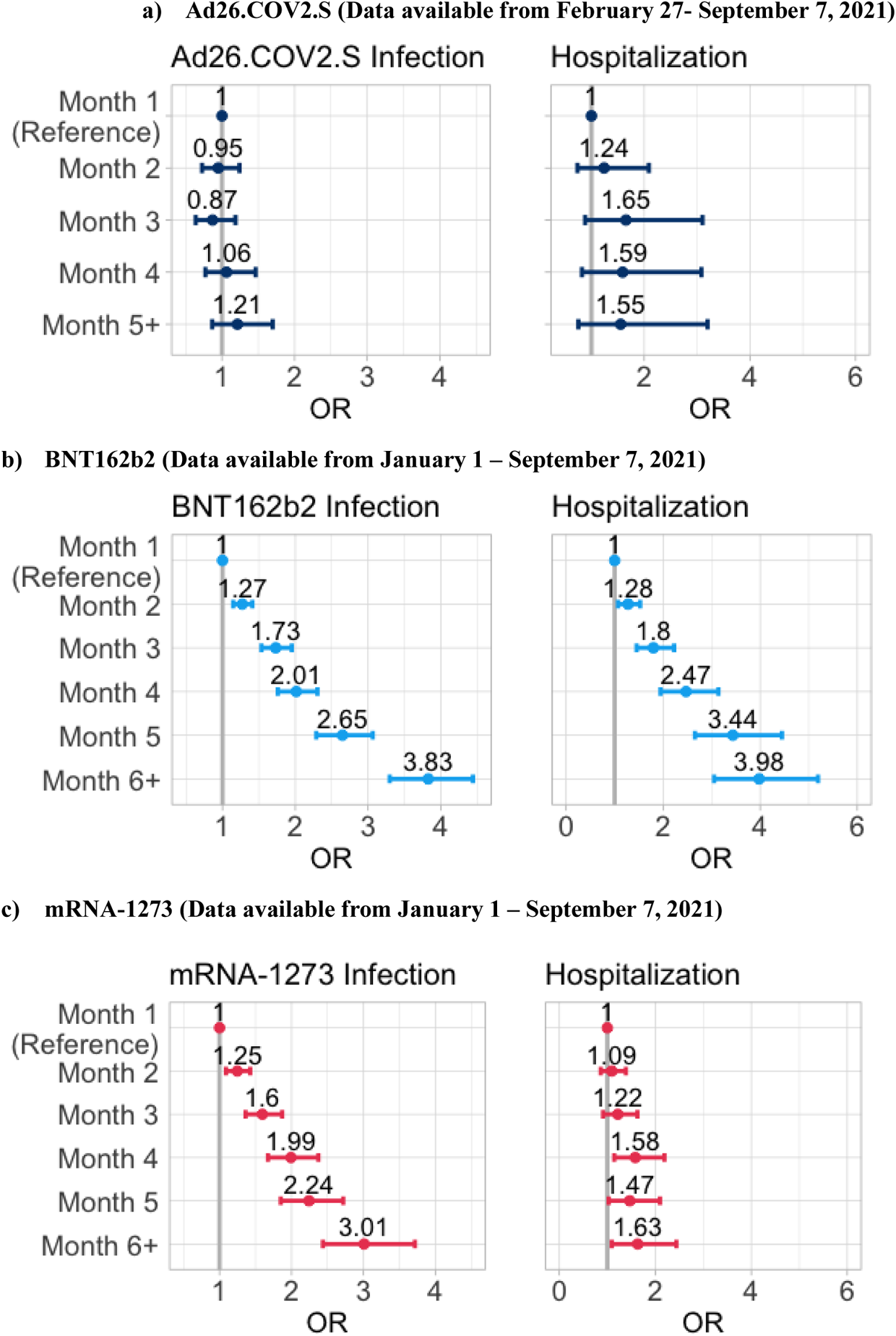
OR and 95% CI assessing durability of baseline vaccine protection against infections and hospitalizations in persons with comorbidity score greater than or equal to 2. CI=confidence interval. OR=odds ratio.

**Fig. S6.**
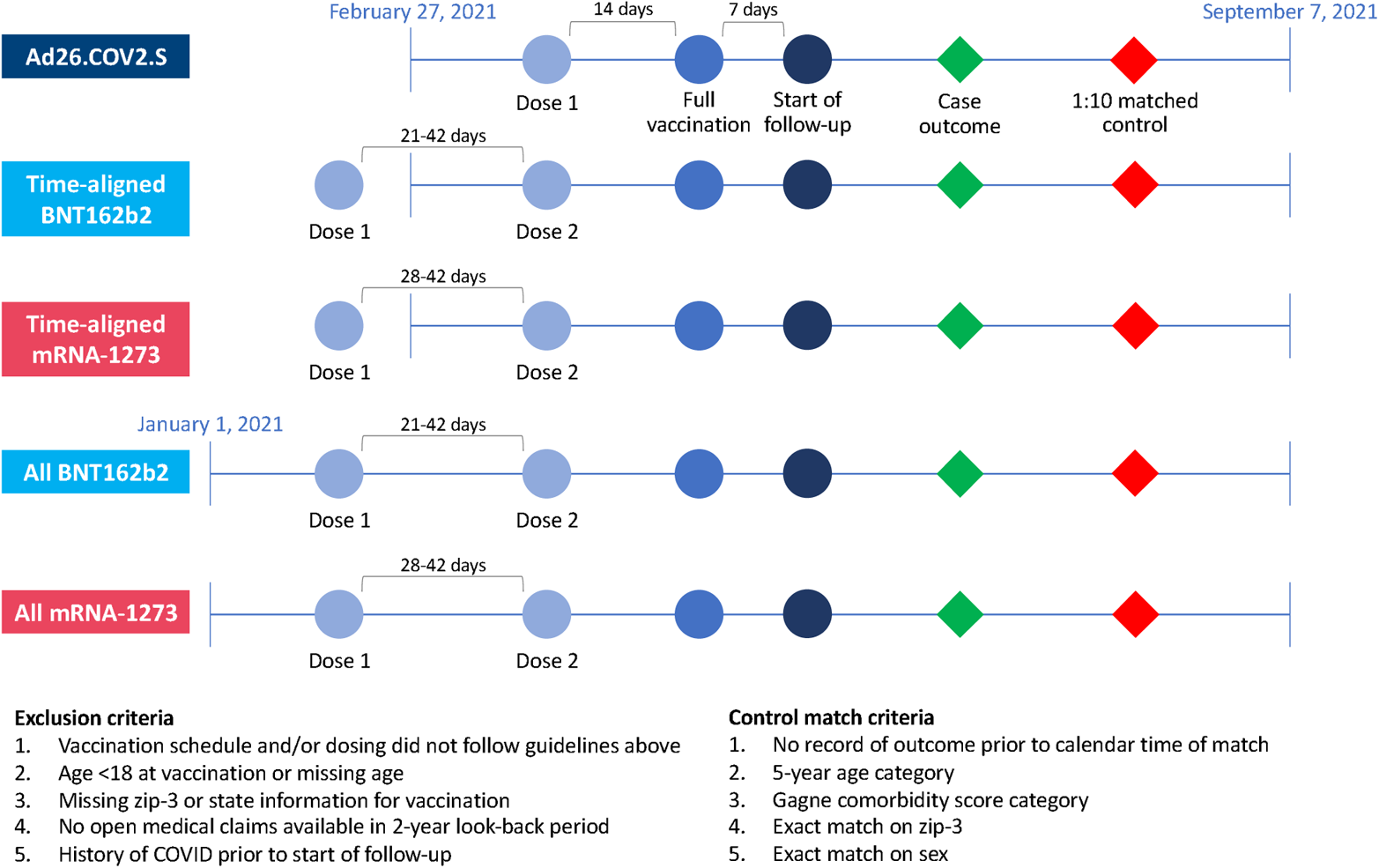
Schematic of the study design. Note: Cases and controls were matched on outcome calendar time. Follow-up time was determined by looking back to the start of follow-up, which varied for cases and matched controls.

**Table S1.** Number of cases and matched controls by vaccine and outcome.

| **Vaccine** | **Outcome** | **Cases**<br>**(N)** | **Matched controls**<br>**(N)** |
| --- | --- | --- | --- |
| Ad26.COV2.S | Infection | 7945 | 67,771 |
|  | Hospitalization | 823 | 5991 |
|  | ICU admission | 82 | 595 |
| All BNT162b2 | Infection | 27,797 | 261,200 |
|  | Hospitalization | 4525 | 42,233 |
|  | ICU admission | 320 | 2989 |
| BNT162b2<br>(for primary vaccine regimens<br>using a subset of data available<br>from February 27, 2021 –<br>September 7, 2021) | Infection | 17,774 | 167,218 |
|  | Hospitalization | 1736 | 15,223 |
|  | ICU admission | 160 | 1410 |
| All mRNA-1273 | Infection | 15,694 | 148,709 |
|  | Hospitalization | 2070 | 19,176 |
|  | ICU admission | 212 | 2057 |
| mRNA-1273<br>(for primary vaccine regimens<br>using a subset of data available<br>from February 27, 2021 –<br>September 7, 2021) | Infection | 11,883 | 112,583 |
|  | Hospitalization | 1336 | 12,112 |
|  | ICU admission | 134 | 1306 |

ICU, intensive care unit.

**Table S2.** Characteristics of cases receiving BNT162b2 in January – February 27, 2021 and February 27 – September 7, 2021.

|  | Infections |  | Hospitalizations |  |
| --- | --- | --- | --- | --- |
|  | Cohort excluded from the time aligned analysis (N=10,051) | Time aligned cohort (N=17,774) | Cohort excluded from the time aligned analysis (N=2791) | Time aligned cohort (N=1736) |
| **Age** |  |  |  |  |
| Mean (SD) | 72.1 (19.0) | 51.3 (18.2) | 80.3 (13.8) | 67.7 (17.1) |
| Median [Min, Max] | 75.0 [18.0, 94.0] | 52.0 [18.0, 94.0] | 81.0 [19.0, 94.0] | 69.0 [18.0, 94.0] |
| Missing | 0 (0%) | 1 (0.0%) | 0 (0%) | 0 (0%) |
| **Age group** |  |  |  |  |
| 18-24 | 198 (2.0%) | 1517 (8.5%) | 3 (0.1%) | 31 (1.8%) |
| 25-29 | 156 (1.6%) | 1045 (5.9%) | 8 (0.3%) | 20 (1.2%) |
| 30-34 | 210 (2.1%) | 1275 (7.2%) | 13 (0.5%) | 39 (2.2%) |
| 35-39 | 243 (2.4%) | 1361 (7.7%) | 16 (0.6%) | 38 (2.2%) |
| 40-44 | 271 (2.7%) | 1373 (7.7%) | 17 (0.6%) | 37 (2.1%) |
| 45-49 | 283 (2.8%) | 1440 (8.1%) | 30 (1.1%) | 63 (3.6%) |
| 50-54 | 394 (3.9%) | 1625 (9.1%) | 44 (1.6%) | 114 (6.6%) |
| 55-59 | 495 (4.9%) | 1877 (10.6%) | 82 (2.9%) | 143 (8.2%) |
| 60-64 | 639 (6.4%) | 1965 (11.1%) | 130 (4.7%) | 194 (11.2%) |
| 65-69 | 872 (8.7%) | 1462 (8.2%) | 199 (7.1%) | 221 (12.7%) |
| 70-74 | 1218 (12.1%) | 1131 (6.4%) | 308 (11.0%) | 227 (13.1%) |
| 75-79 | 1205 (12.0%) | 715 (4.0%) | 395 (14.2%) | 188 (10.8%) |
| 80-84 | 984 (9.8%) | 423 (2.4%) | 349 (12.5%) | 157 (9.0%) |
| 85+ | 2883 (28.7%) | 565 (3.2%) | 1197 (42.9%) | 264 (15.2%) |
| **Sex** |  |  |  |  |
| Female | 6268 (62.4%) | 10,380 (58.4%) | 1637 (58.7%) | 882 (50.8%) |
| Male | 3783 (37.6%) | 7394 (41.6%) | 1154 (41.3%) | 854 (49.2%) |
| **Gagne** |  |  |  |  |
| Mean (SD) | 2.21 (2.82) | 0.771 (1.82) | 3.04 (2.96) | 2.44 (3.08) |
| Median [Min, Max] | 1.00 [-1.00, 18.0] | 0 [-2.00, 18.0] | 2.00 [-1.00, 17.0] | 1.00 [-1.00, 18.0] |
| **Gagne group** |  |  |  |  |
| <2 | 5304 (52.8%) | 14,528 (81.7%) | 1050 (37.6%) | 875 (50.4%) |
| 2-3 | 2124 (21.1%) | 1961 (11.0%) | 711 (25.5%) | 375 (21.6%) |
| 4-5 | 1240 (12.3%) | 671 (3.8%) | 475 (17.0%) | 206 (11.9%) |
| 6+ | 1383 (13.8%) | 614 (3.5%) | 555 (19.9%) | 280 (16.1%) |
| **Medicaid** |  |  |  |  |
| Yes | 2267 (22.6%) | 2502 (14.1%) | 793 (28.4%) | 336 (19.4%) |
| No | 7784 (77.4%) | 15,272 (85.9%) | 1998 (71.6%) | 1400 (80.6%) |

SD, standard deviation.

